# Transfer functions: learning about a lagged exposure-outcome association in time-series data

**DOI:** 10.1101/2021.09.30.21264361

**Authors:** Hiroshi Mamiya, Alexandra M. Schmidt, Erica E. M. Moodie, David L. Buckeridge

## Abstract

Many population exposures in time-series analysis, including food marketing, exhibit a time-lagged association with population health outcomes such as food purchasing. A common approach to measuring patterns of associations over different time lags relies on a finite-lag model, which requires correct specification of the maximum duration over which the lagged association extends. However, the maximum lag is frequently unknown due to the lack of substantive knowledge or the geographic variation of lag length. We describe a time-series analytical approach based on an infinite lag specification under a transfer function model that avoids the specification of an arbitrary maximum lag length. We demonstrate its application to estimate the lagged exposure-outcome association in food environmental research: display promotion of sugary beverages with lagged sales.

## Introduction

Many population exposures have time-lagged association with population health outcomes, cumulating their impact over time^1,2^. An often-overlooked aspect of public health nutrition, for example, is the impact of unhealthy food promotions which frequently exhibit time-lagged associations with outcomes such as unhealthy food purchasing. In other words, exposures show a persistent association with outcomes in future times periods. Distributed lag models capture such lagged patterns by incorporating a series of lagged values of an exposure, whose coefficients typically are characterized by a functional structure such as low-order polynomials or splines ^1,2^. Aside from the specification of the degree of polynomials, these models require a priori knowledge about the maximum number of lags (hereafter referred to as lag length), since incorrect specification of the lag length may fail to capture the lag structure in terms of shape and magnitude ^3–5^. However, the plausible lag length for a given exposure-outcome association is often unknown due to the lack of substantive knowledge and, in the case of food promotions as an exposure, the lag length may vary widely across geography and food categories of varying shelf-life, such as soda vs. yoghurts^3,4,6^.

In this brief report, we describe a distributed lag model with an infinite lag specification under a transfer function as an approach to obviate the need for a priori specification of lag length. We motivate this with an analysis which aims to capture the lagged association between an exposure related to an obesogenic food environment (weekly time-series of within-store display promotion of sugary beverages) on the purchasing ultra-processed food (weekly sales of such beverages).

## Methods

We assume that measurements are available at discrete and equally spaced intervals, and focus on a widely used and interpretable specification of the transfer function called the Koyck decay^7^, a monotonic decrease of association over time capturing the diminishing association of an exposure such as media advertising with food purchasing^4^.

A transfer function is a parsimonious specification of lagged terms and uses a structural variable, denoted *E*_*t*_ below, to summarize the current (at time *t*) and cumulative association (up to time *t*) between the exposure variable *x* and the outcome *y*. Using a decay parameter *λ* and a lag index *h*, monotonically decreasing associations are represented as;

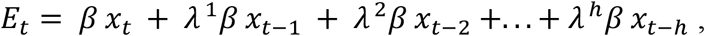

which recursively reduces to

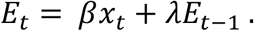

The coefficient *β* captures the *immediate* association at time *t*, while the coefficient *λ* captures the extent of the lag: a value closer to 1 implies a more persistent lag, while a value nearer to zero indicates a shorter lag. When the structure *E*_*t*_ is added to a regression model, learning of a lagged association is accomplished by estimating *λ* and *β*.

An interpretation of a lagged association combining these two parameters is called an *impulse response function*. This represents the response (change) of *y*_*t*+0_ + *y*_*t*+1_ + *y*_*t*+2_ + … + *y*_*t*+*h*_ to impulse (one-unit increase of *x* at time *t* only), while all other variables at time *t* or earlier are fixed constant ^8^. For the Koyck lag, the impulse response function takes the form *βλ*^0^ + *βλ*^1^ + *βλ*^2^ + … + *βλ* ^*h*^ as visualized in Supplementary Figure S1a-b and Supplementary Appendix S1. Imposing the constraint −1 < λ < 1 will ensure that this function decays to zero as *h* → ∞.

The general specification of the transfer function governing the shape of lag is as follows;

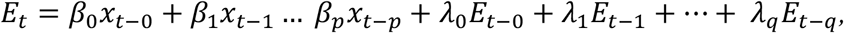

where the Koyck decay is defined by *p* = 0, *q* = 1. More complex shapes can be specified by higher order transfer functions, i.e., using higher values of *p* and *q*, as long as a large number of *p* does not lead to collinearity across variables *x*_*t*−*p*_. Possible shapes include a peak after several lags followed by decay. Further discussion of transfer functions is provided elsewhere.^9–12^

Clearly, there is no need to provide a lag length in this infinite lag specification, but a priori knowledge about plausible shapes of the lag is needed, along with constraints on the value of *λ*: in case of the Koyck lag, 0 < λ < 1 ensures monotonic decay, while −1 < *λ* < 0 leads to an oscillating lag (Supplementary Figure 1c). The selection of candidate shapes may rely on model selection, for example guided by metrics for model fit (an information criterion) or forecasting error. This is in contrast to a commonly used finite lag model, where the shape of the lag is learned from data and the plausible (or arbitrary) value of lag length is fixed in advance by the investigator.

### Application

The exposure of interest is the weekly within-store display promotion of an obesogenic food product (sugary beverages) that potentially exhibits time-lagged association with the outcome, purchasing quantity of sugary beverages in the same store. Display promotion is the temporary placement of selected items in prominent locations and is shown to have a positive association with the sales of ultra-processed (‘junk’) food, such as sugar sweetened beverages. This exposure is thus increasingly recognized as the policy target to improve population diets^13^. Our food category of interest is sugar sweetened (not plain) drinkable yogurt, which is a hidden and important source of dietary sugar, in particular among children ^14,15^. The weekly status of the proportion of display-promoted sweetened yogurt items (continuous exposure) and the weekly aggregated sum of the sales quantity of sugar-sweetened drinkable yogurt items (continuous outcome) were generated by a single large supermarket in Montreal, Canada over *n*=311 weeks. A detailed description of the promotion exposure and outcome are provided in Supplementary Appendix S2.

Weekly sales were modelled as a function of the structural variable *E*_*t*_, covariates, a seasonal term, and an intercept. We selected the Koyck lag specification with *λ* bounded to the interval [0,1], given that the promotion is expected to have a monotonically decaying association with purchasing due to temporarily decreasing awareness among exposed consumers^4^. The time-series regression used in this study is a dynamic linear model, which allows regression parameters to vary smoothly over time^10,16^. The model was fit under the Bayesian framework. The details of the dynamic linear model, model specification, and prior specification are provided in Supplementary Appendix S3. The parameters of interest are *β* and *λ*, from which the impulse response function was estimated. Code to perform the analysis is provided at https://github.com/hiroshimamiya/promotionLag/blob/main/discountLag_KoyckTransfer.stan

## Results

The estimated posterior mean of *β* was 0.68 (95% Credible Interval [CI]: 0.39-0.96), an association implying exp(0.68) higher absolute (i.e., non-log) quantity of sugar-sweetened drinkable yogurt at week *t* if all yogurt items were display promoted in that week. The posterior summary of the decay coefficient *λ* was modestly strong: 0.47 (95% CI 0.20-0.72), as shown by the distinct lag in the estimated impulse response function (Figure 1). The posterior median of the excess quantity of sales associated with lag alone on the absolute (*non-log*) scale was 11,890 (95% CI 2,594-32,051) servings for the entire 6 years period, while that of excess sales related to the immediate association alone was 13,506 (95% CI: 7,726-19,750). An autocorrelation function plot of residuals over time suggests that temporal autocorrelation in the fitted model was largely absent (Supplementary Figure S4).

**Figure 1:**
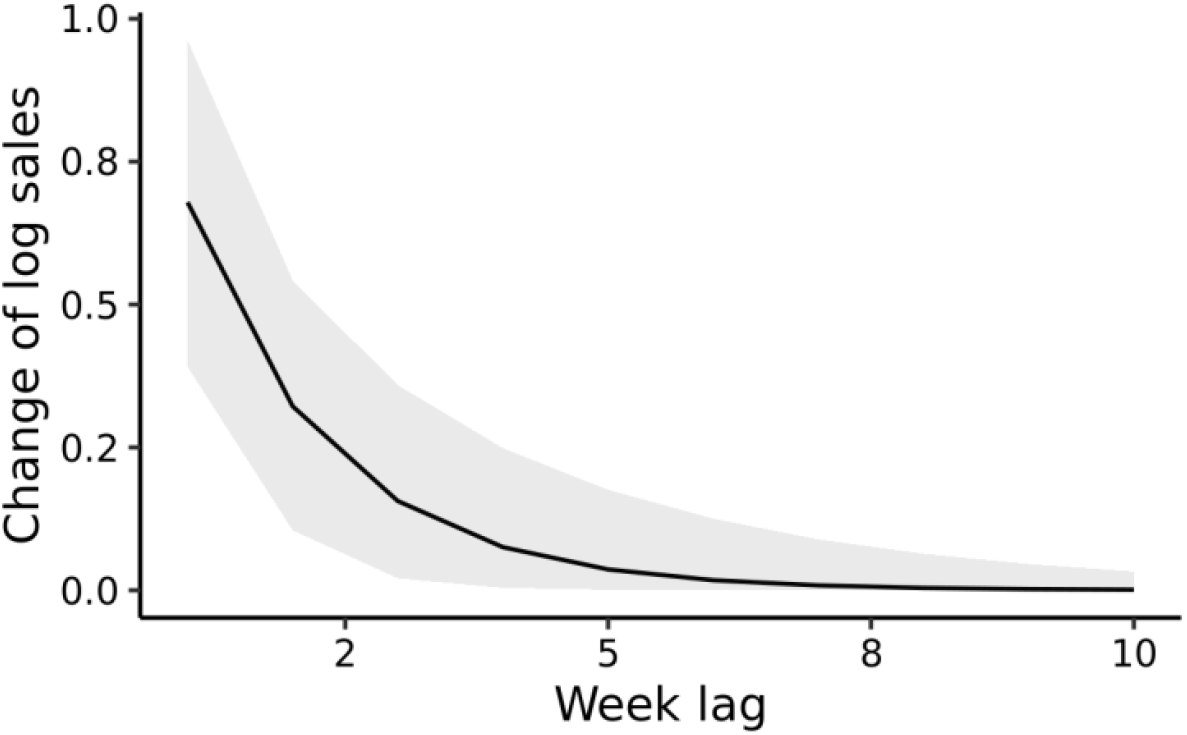
The estimated impulse response function of display promotion on the natural log sales of sugar-sweetened drinkable yogurt, based on the lag parameter *β* and *λ* learned from the time-series of sales data from a single store in Montreal (*n*=311 weeks, 2008-2013). The grey band indicates pointwise 95% posterior credible interval. The immediate association is displayed at lag 0 and is 0.68 (95% Posterior Credible Interval: 0.39-0.96), indicating that the immediate impact of display promotion is a doubling of sales, since exp(0.68) = 1.97.

## Discussion

Time-lagged associations between exposures and outcomes are ubiquitous and, therefore, are important components to decompose in time-series studies. We described the transfer function as an approach to capture lagged association when the lag length is unknown, with an application in food environment research.

The transfer function approach is particularly useful when lag length may vary across many food categories and/or store-neighborhood socio-economic status, the latter being particularly relevant when investigating the role of food promotions in driving geographic and socio-economic disparities of diets. The transfer function can also be used in climate-related environmental time-series analysis to investigate spatially heterogeneous persistence of lagged association between temperature and arbovirus incidence^17^. The relatively simple Koyck lag function may also be used to capture the monotonically decaying lagged association of high ambient temperature with mortality and morbidity, although there may be a need to account for mortality displacement in some geographic regions^6^.

The added value of the proposed approach in estimating a valid lag structure relies on researcher’s prior knowledge of plausible lag shapes i.e., values of *p* and *q* in the transfer function. Thus, when such knowledge is lacking, existing finite lag models using polynomial or basis function will allow data-driven estimation of the approximate shape of lag, conditional on the availability of theoretical or empirical evidence about lag length. However, the finite lag approach requires other knowledge, specifically the lag length.

Limitations of the current approach include challenges in selecting the most appropriate shape of transfer function, for example when competing shapes show similar model fit and thus the underlying process generating lagged association is undetermined.

Modeling exposure-lag association using a transfer function addresses one of the current methodological gaps in epidemiological time-series analysis, which is the potential bias in lagged association due to incorrect specification of lag length in finite lag model. The proposed approach shown here can be readily implemented using time-series regression and codes provided.

## Data Availability

Scanner transaction data from retail food outlets are collected in many nations by the Nielsen company (https://www.nielsen.com/ca/en/solutions/measurement/retail-measurement/). The data are available by commercial agreement or through affiliated academic institutions that maintain access to these data for research use.

