## Supplementary Material for "Transfer functions: learning about a lagged exposure-outcome association in time-series data"

### Supplementary Appendix S1: Impulse response of Koyck function

Supplementary Figure S1 below provides examples of Koyck decay functions generated by the coefficient of the immediate association $\beta$ and the coefficient of decay $\lambda$ as described in the main document. Exposure in this plot represents a shock in time-series (i.e., a “one time” impulse exposure); its value is increased by one unit at lag 0, then set to zero at subsequent lag points. The y-axis represents the change in the association between the exposure and outcome, such as a relative risk for a count outcome (i.e., exponentiated regression coefficient) or a change in a continuous outcome, such as sales. Note that not constraining the value of $\lambda$ to [-1,1] results in non-stationary time-series with the lagged association asymptotically decaying to non-zero.

The lagged associations appearing in Supplementary Figure S1 represent an impulse response function, which is a polynomial function representing the expected change of

$y_{t+0} + y_{t+1}+y_{t+2} + ...+ y_{t+h}$. The immediate (not lagged) association at the time of exposure at lag zero (*x=0* in the figures below) is $\beta\lambda^{0}=$ $\beta$.

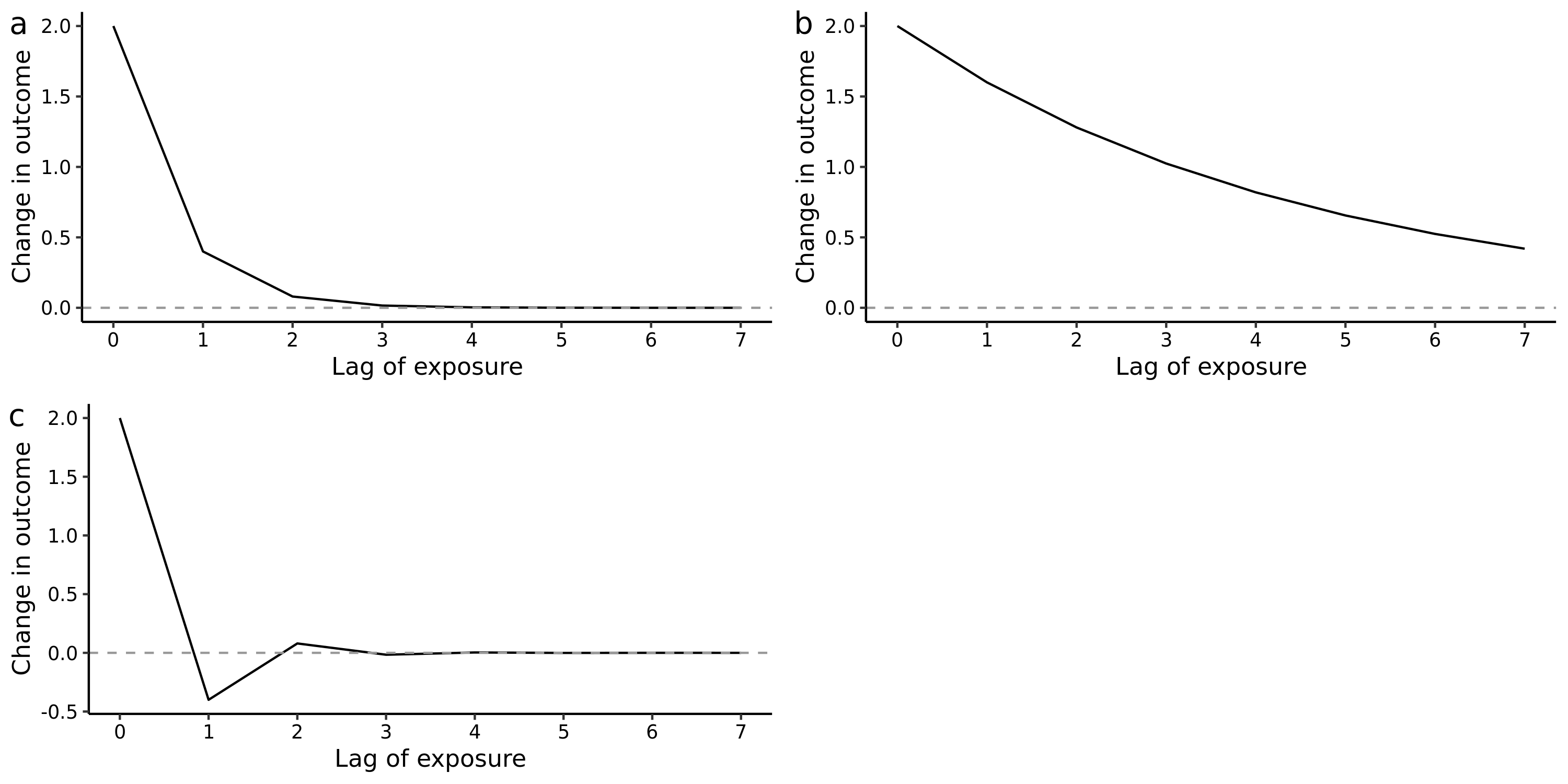

**Supplementary Figure S1**: Impulse response function of the Koyck transfer function, with the speed and extent of the decay controlled by the value of lag parameter $\lambda$: a) a monotonic and weak decay returning to the baseline with a short lag ($\lambda=0.2)$: b) a more persistent lag ($\lambda=0.8)$: and c) an oscillating function when the value is negative ($\lambda=-0.2$).

### Supplementary Appendix S2: Definition of exposure and outcome

The food category of interest is sugar-sweetened drinkable yogurt, which is classified as ultra-processed food^1^ and considered to be the source of excess sugar intake, along with other sugar-sweetened beverages such as soda^2^. We excluded plain yogurt items that contain intrinsic sugar only (i.e., fructose) were excluded, as they are classified as minimally processed food and thus non-sugar-sweetened. Weekly beverage transaction and promotion data in this study were purchased from a global marketing company, Nielsen, that collects scanned electronic transaction records from chain retail food outlets^3^.

The outcome is the weekly sum of the sales of sugar-sweetened drinkable yogurt items sold in a large supermarket in Montreal, Canada between January 2008 and December 2013 (n=311 weeks). There were 29 distinct sugar-sweetened yogurt items that were display promoted at least once during the study period. Sold quantities of individual items were summed at each week as a single time-series and natural log-transformed to approximately follow a normal distribution. The descriptive plot of sales is provided in Supplementary Figure S2 below.

Exposure is the aggregated weekly display promotion status of sugar-sweetened drinkable yogurt items. Display promotion is the form of in-store marketing that temporarily places selected food items to prominent locations, such as the store entrance, at the end of aisle, or at the checkout aisle in order to increase awareness of promoted items^4^. This promotion often (but not always) occurs with other forms of promotions, such as price discounting and flyers - see the section below describing covariates in the time-series regression model below. The weekly time-series of display promotion was defined as the proportion of displayed items in that week among sweetened drinkable yogurt items (thus ranging from 0 to 1).

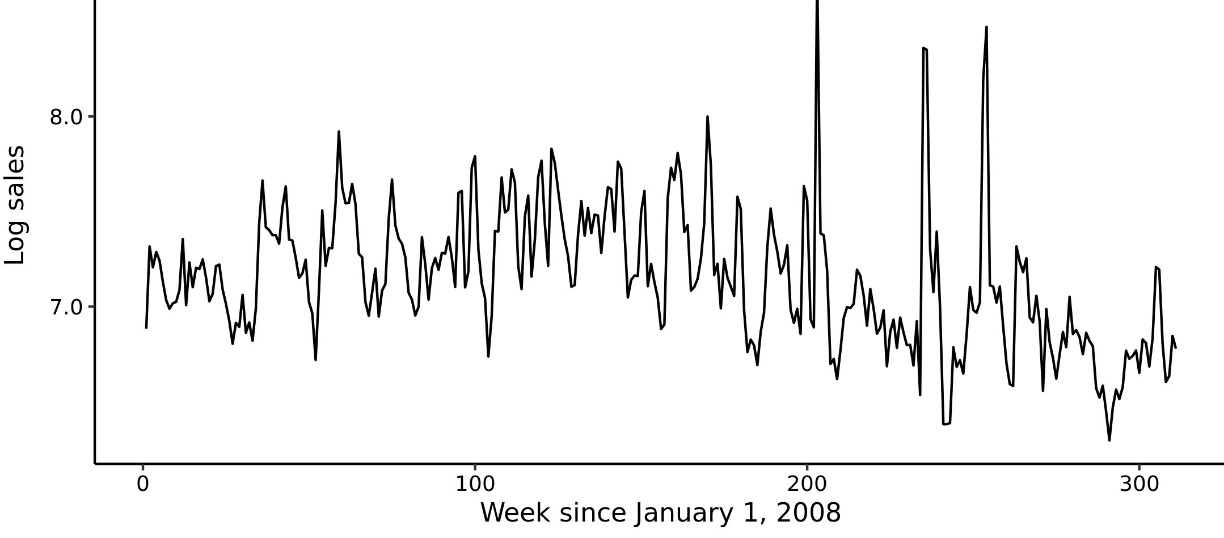

Supplementary Figure S2. Natural log-transformed weekly sales of 29 yogurt items in a single supermarket, Montreal, Canada between January 2008 and December 2013.

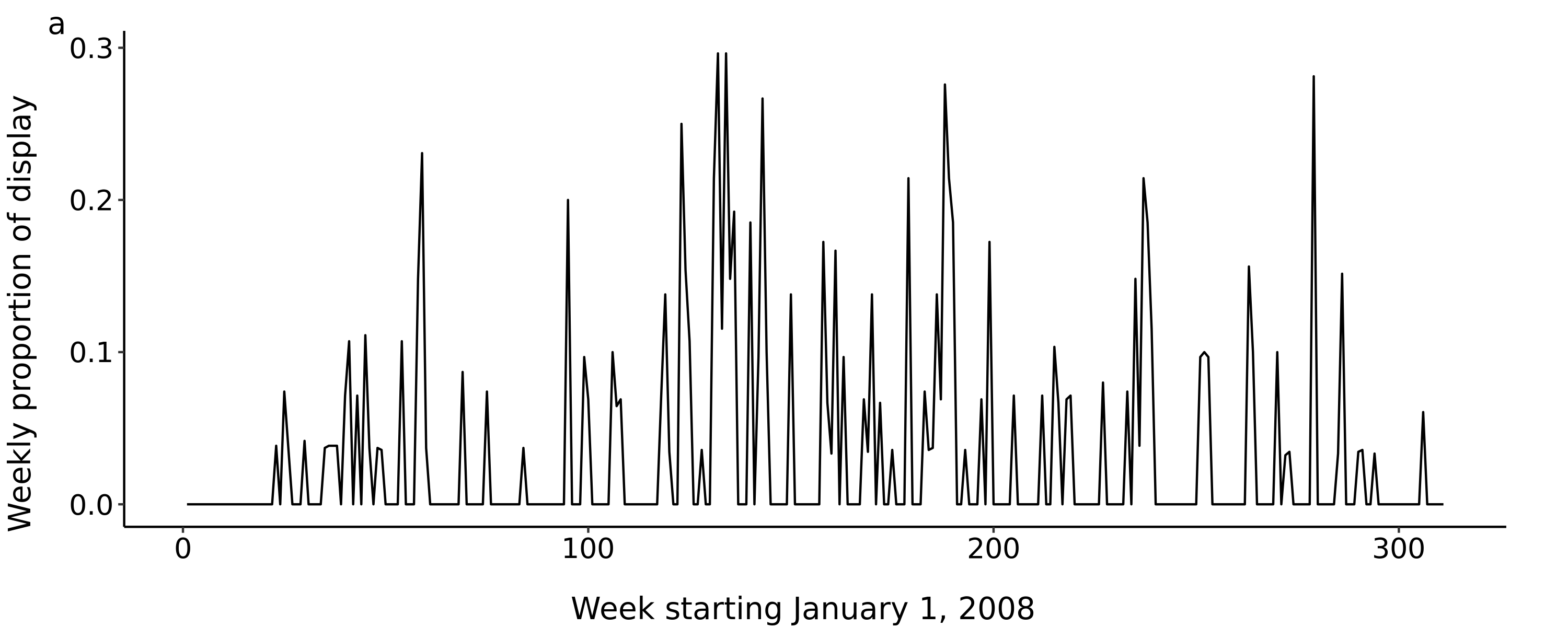

Supplementary Figure S3. Weekly proportion of 29 yogurt items in a single supermarket, Montreal, Canada between January 2008 and December 2013.

### Supplementary Appendix S3: Time-series regression

The structural variable $E_{t}$, defined in the main article, was added to a dynamic linear model, which allows regression coefficients to evolve smoothly (‘dynamic’) over time.^5,6^ The outcome in this study, denoted $y_{t}$, is the natural log-transformed quantity of sales at week *t* and assumed to be normally distributed as $y_{t} \sim Normal(\mu_{t}, \epsilon_{t}).$The mean is the linear combination of the parameters as shown below:

$$\mu_{t}=\alpha_{t}+S_{t}+\gamma C_{t}+E_{t},$$

where $\alpha_{t}$ is a time-varying (dynamic) intercept capturing trends and local fluctuation of outcome, $S_{t}$ is seasonal terms with a periodic cycle, and $C_{t}$ represents a vector of covariates with the corresponding coefficient vector $\gamma$. The error term, $\epsilon_{t},$ is a normally distributed noise with time-fixed variance parameter as $\epsilon_{t} \sim Normal(0, \sigma_{\epsilon})$.

The local shift of the intercept at time *t*, $\alpha_{t}$ is determined by its previous value, $\alpha_{t-1}$, and random noise term $\epsilon_{alpha}$; thus $\alpha_{t}=\alpha_{t-1}+\epsilon_{alpha}$ accounts for the temporal correlation of the outcome. The noise term, $\epsilon_{alpha}$, follows a zero-mean independent normal distribution $\epsilon_{alpha} \sim Normal(0, \sigma_{\alpha})$ with the smoothness controlled by $\sigma_{\alpha}$.

The seasonal component $S_{t}$ is a harmonic representation of time with a sinusoidal wave. The periodicity is defined as $2\pi t\omega$, where $t=\{1,2,..., 311\}$ as the week indicator, $\omega$ = 52.2 as the scalar representing the cycle (number of weeks in a year) and $\pi$ = $3.1415...$. Regression coefficients, $\gamma_{Cos}$ and $\gamma_{Sine}$, capture the amplitude of the seasonal association as follows:

$S_{t}=\gamma_{Cos}Cos\left( 2\pi t\omega\right)+\gamma_{Sine}Sine\left( 2\pi t\omega\right)$.

Note that $\gamma_{cos}$ and $\gamma_{sine}$ are set to be time-fixed in this example (i.e., not dynamic).

Finally, $C_{t}$ is a vector of weekly-varying covariates known to temporarily correlate with display promotion and sales, and $\gamma$ is the corresponding time-fixed regression coefficients. The covariates include weekly price discounting as defined in our previous study^7^, consumer price index as the indicator of inflation, flyer promotion defined in the same manner as display promotion, and the binary indicator of week containing provincial and national statutory holiday. Selection of the covariates were guided by Watanabe-Akaike Information Criterion that indicates a better model fit when its value is lower.^8^

In our study, the parameters above were estimated by Hamiltonian Monte Carlo under the Bayesian Framework^5^ using the rstan library in R software^9^. We ran 3,000 iterations as a burn-in sample, and the following 30,000 iterations were used to simulate from the posterior distribution of the parameters. Codes are available in :

<https://github.com/hiroshimamiya/promotionLag/blob/main/discountLag_KoyckTransfer.stan>.

Convergence of the Markov Chain Monte Carlo was inspected visually on the trace plots of the parameters from 3 independent chains.

The prior probability of the error terms $\varepsilon$ was specified as a zero-mean non-informative (i.e., diffuse) Cauchy distribution with the variance $\varepsilon^{+} \sim Cauchy(0, 5^{2})$ as previously suggested^10^. The superscript ^+^ indicates that the distribution is constrained to take a positive value (i.e., constrained to the positive half of the Cauchy distribution). To investigate whether posterior distribution is affected by the scale (diffuseness) of the prior distribution, we also ran model with a larger and smaller value of variance for the error term, $\varepsilon^{+} \sim Cauchy(0, {10}^{2})$ and $\varepsilon^{+} \sim Cauchy(0, 3^{2})$, respectively, which generated nearly identical posterior distribution. We also used a positive-constrained normal distribution to examine posterior sensitivity with the same values of the scaling parameter as above, which led to nearly identical results.

The prior probability of the regression coefficients, including the season coefficients was specified as independent non-informative normal distributions $\gamma, \gamma_{cos}{,\gamma}_{sine}, \beta\sim Normal(0,5^{2})$. Again, we experimented with a larger value of the variance, $Normal(0,{10}^{2})$, and a smaller value, $Normal(0,3^{2})$, resulting in nearly identical posterior distributions. For the time-varying intercept, $\alpha_{t}$, the prior probability of the initial value at $t = 1$ was $\alpha_{1}\sim Normal(0,5^{2})$, which was subsequently allowed to evolve with a random noise $\epsilon_{alpha} \sim N(0,1)$. The lag parameter was assigned a non-informative uniform distribution $\lambda\sim Uniform(0,1)$*.*

While the outcome in this application was considered normally distributed, the model is generalizable to outcome variables in the exponential family, including commonly modelled count outcomes such as mortality and morbidity in environmental time-series analysis (Poisson distribution) with overdispersion parameters^5,6^. As well, non-Bayesian approach to estimate the correlated dynamic regression coefficients ($\alpha_{t}$ in our study) is also possible using established formulas via the Kalman filter and smoother.^6^

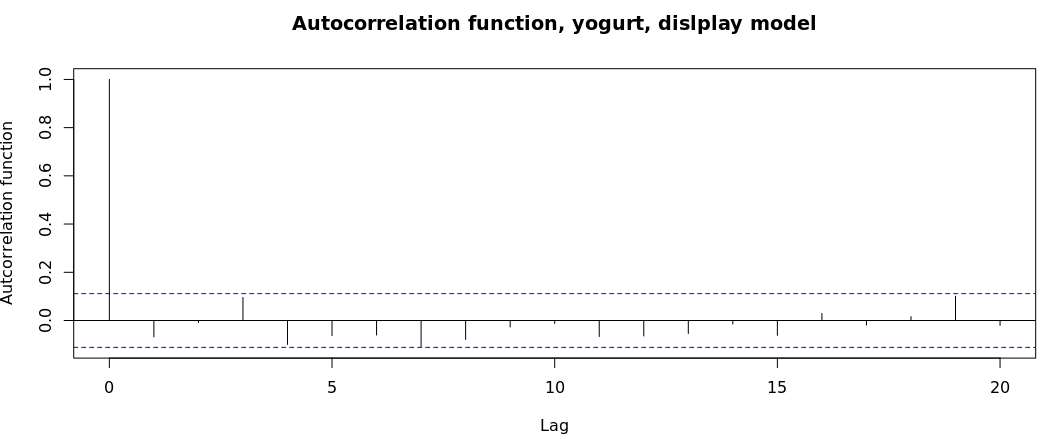

Supplementary Figure S4. Autocorrelation function of residuals.
